# FLOT versus SOX Neoadjuvant Chemotherapy: First Prospective Survival Comparison in Locally Advanced Gastric Cancer

**DOI:** 10.1101/2025.06.07.25329179

**Authors:** Birendra Kumar Sah, Chen Li, Zhenggang Zhu

## Abstract

**Background:** Both FLOT and SOX neoadjuvant regimens are widely used for locally advanced gastric cancer; however, direct head-to-head survival data to guide optimal treatment selection are lacking.

**Methods:** We conducted an open-label, randomized, phase 2 trial (NCT03636893) at a single center in China. Patients with locally advanced gastric cancer (cT3-4b, cN1-3, cM0) were randomized to receive neoadjuvant FLOT (four cycles) or SOX (three cycles) before D2 gastrectomy. We report the long-term survival outcomes with 5-year follow-up in 74 randomized patients through May 2025.

**Results:** The first prospective comparison demonstrated remarkable outcomes for both regimens. With median follow-up of 65.7 months, median overall survival exceeded 5 years in both groups: 61.5 months (95% CI: not reached) for FLOT versus 67.8 months (95% CI: 25.7-109.9) for SOX, with no significant difference (HR 1.101, 95% CI: 0.595-2.036, p=0.759). Disease-free survival was equivalent (23.0 vs 25.5 months, HR 1.060, p=0.842). Clinicopathological factors proved to be more prognostic than regimen choice: complete/subtotal tumor regression achieved 80.5-month survival versus 47.6 months for partial response (p=0.017), whereas gastrectomy type emerged as the strongest independent predictor (HR 3.619 for total vs. partial gastrectomy, p=0.010). Both regimens demonstrated favorable safety profiles with manageable toxicity.

**Conclusions:** This study establishes equivalent long-term survival between the FLOT and SOX regimens. With both achieving 5-year survival, treatment selection should prioritize patient factors, institutional experience, and practical considerations, rather than expected survival differences. Optimizing the pathological response and surgical approach appears more critical than specific regimen choice.

**Funding:** This research received no specific grant funding.

## INTRODUCTION

For millions of patients diagnosed with locally advanced gastric cancer (LAGC) worldwide, selecting an optimal neoadjuvant chemotherapy regimen represents one of the most consequential treatment decisions in modern oncology. Gastric cancer remains the fourth most common malignancy and second leading cause of cancer-related mortality globally, with particularly high incidence rates in East Asian countries.^1^

The perioperative chemotherapy landscape transformed dramatically following landmark trials demonstrating clear survival benefits over surgery alone.^2–5^ The German FLOT4 trial revolutionized treatment by establishing FLOT (docetaxel, oxaliplatin, leucovorin, and 5- fluorouracil) as superior to ECF/ECX regimens, with significant improvements in both pathological response and overall survival.^6,7^ This led to FLOT adoption as the preferred regimen in Western guidelines.^8,9^

However, a parallel evolution occurred in East Asian countries, where different patient populations and healthcare systems favored oral fluoropyrimidine-based approaches. The SOX regimen (S-1 plus oxaliplatin) emerged as a compelling alternative, with large-scale Chinese trials (RESOLVE and RESONANCE) suggesting comparable efficacy to triplet regimens while maintaining favorable tolerability.^10,11^ SOX offers practical advantages: reduced infusion time, fewer cycles, and potentially lower rates of severe neutropenia.^12^

The treatment landscape continues evolving with immunotherapy integration. The phase 3 DRAGON IV trial demonstrated that adding camrelizumab and rivoceranib to SOX significantly improved pathologic complete response.^13^

Despite widespread clinical use of both regimens, a critical knowledge gap persists: direct comparative data regarding long-term survival outcomes remain limited. Most published studies have been single-arm phase 2 trials or retrospective analyses, with few prospective head-to-head comparisons available.

We previously reported comparable pathological response rates between FLOT and SOX in the Dragon III trial (20.0% vs. 32.4% complete/subtotal tumor regression, p=0.289).^14^ However, pathological response may not always translate directly into survival benefit, requiring validation with long-term survival data.

The primary objective was to compare long-term survival outcomes between neoadjuvant FLOT and SOX regimens in patients with LAGC using extended follow-up data from the Dragon III trial. Secondary objectives included analyzing disease recurrence sites, identifying prognostic factors, and exploring relationships between pathological response and clinical outcomes.

## METHODS

### Study Design and Setting

The Dragon III trial was an investigator-initiated, phase 2, open-label, randomized controlled trial comparing neoadjuvant FLOT and SOX regimens for patients with locally advanced gastric cancer. The trial design, eligibility criteria, and treatment protocols have been published previously.^14^ This manuscript reports the pre-specified secondary endpoints of overall survival (OS) and disease-free survival (DFS) with extended follow-up through May 8, 2025. The CONSORT flow diagram detailing patient enrollment, allocation, follow-up, and analysis was published in our initial report^14^ and is not reproduced here. The trial was prospectively registered at ClinicalTrials.gov (NCT03636893) before patient enrollment.

The study was conducted at Ruijin Hospital, Shanghai Jiao Tong University School of Medicine between August 2018 and March 2020. The Institutional Review Board of Ruijin Hospital approved the study protocol, including long-term follow-up plans. All patients provided written informed consent for participation and long-term follow-up.

### Patient Selection and Eligibility

#### Inclusion criteria were designed to select patients with resectable locally advanced disease

Patients aged 18-80 years with histologically confirmed adenocarcinoma of the stomach or esophagogastric junction were eligible if they had clinical stage cT3-cT4b, cN1-cN3, cM0 disease. Key inclusion criteria included adequate organ function, ECOG performance status ≤2, and written informed consent. The detailed inclusion and exclusion criteria have been published elsewhere.^14^ The baseline characteristics of the randomized patients are shown in Table 1.

**Table 1:** Patient Demographics and Baseline Characteristics.

| **Characteristic** | **FLOT Group (n=40)** | **SOX Group (n=34)** | **p-value** |
| --- | --- | --- | --- |
| Age, median (range), years | 67 (27-76) | 61 (34-80) | 0·239 |
| **Sex - no. (%)** |  |  | 0·428 |
| Male | 29 (72·5) | 21 (61·8) |  |
| Female | 11 (27·5) | 13 (38·2) |  |
| **Clinical TNM Stage - no. (%)** |  |  | 0·365 |
| IIB | 1 (2·5) | 3 (8·8) |  |
| III | 22 (55·0) | 18 (52·9) |  |
| IVA | 17 (42·5) | 13 (38·2) |  |
| **Tumour Location - no. (%)\*** |  |  | 0·622 |
| Proximal | 7 (22·6) | 7 (29·2) |  |
| Proximal body | 3 (9·7) | 1 (4·2) |  |
| Body | 7 (22·6) | 3 (12·5) |  |
| Distal body | 6 (19·4) | 5 (20·8) |  |
| Distal | 8 (25·8) | 6 (25·0) |  |
| Total | 0 | 2 (8·3) |  |
| **Lauren Classification - no. (%)\*** |  |  | 0·389 |
| Intestinal | 18 (58·1) | 13 (54·2) |  |
| Diffuse | 7 (22·6) | 6 (25·0) |  |
| Mixed | 3 (9·7) | 5 (20·8) |  |
| Unclassifiable | 3 (9·7) | 0 |  |

\*Among patients who underwent surgery (FLOT n=31, SOX n=24). TNM=Tumour, Node, Metastasis staging system, 8th edition; FLOT=docetaxel, oxaliplatin, leucovorin, 5-fluorouracil; SOX=S-1 plus oxaliplatin.

### Randomization and Treatment Allocation

#### Randomization was performed using rigorous methodology to ensure balance

Patients were randomized 1:1 to receive either neoadjuvant FLOT (n=40) or SOX (n=34) using simple randomization without stratification. The randomization sequence was generated by an independent statistician and implemented through a central electronic system. Patient screening was conducted by trained clinicians at Ruijin Hospital. Principal investigators evaluated pretreatment assessments and made final enrollment decisions. After confirming eligibility and obtaining informed consent, participants were assigned to treatment groups by the independent statistician using the randomization sequence. Treatment assignments were communicated to clinical teams via telephone or text message. Neither patients nor investigators were blinded to treatment allocation due to the different administration protocols of the regimens.

### Treatment Protocols

#### Both regimens followed established dosing and scheduling

The FLOT regimen consisted of four cycles of docetaxel 50 mg/m^2^, oxaliplatin 85 mg/m^2^, leucovorin 200 mg/m^2^, and 5-fluorouracil 2600 mg/m^2^ every 2 weeks. The SOX regimen consisted of three cycles of oxaliplatin 130 mg/m^2^ on day 1 and oral S-1 80 mg/m^2^ twice daily on days 1-14, repeated every 3 weeks. Surgery was performed 2-4 weeks after completion of neoadjuvant chemotherapy.

#### Surgical procedures were standardized at a high-volume center

All surgical procedures were performed by experienced surgeons at a high-volume center. Standard D2 gastrectomy was performed according to Japanese Gastric Cancer Treatment Guidelines. Adjuvant chemotherapy was administered according to institutional guidelines, with the recommended regimen being a continuation of the preoperative protocol.

### Outcome Definitions and Assessments

#### Primary survival endpoints were rigorously defined

Overall survival (OS) was defined as the time from randomization to death from any cause. Patients alive at the time of analysis were censored at the date of last follow-up contact. Disease-free survival (DFS) was defined as the time from randomization to the first occurrence of local recurrence, regional recurrence, distant metastases, or death from any cause. Disease recurrence was assessed using CT imaging with contrast, upper endoscopy for local recurrence, histological confirmation when feasible, and multidisciplinary team review of imaging findings.

### Pathological response was assessed using validated criteria

Treatment response was assessed using the Becker tumor regression grading system: TRG 1a (complete regression), TRG 1b (subtotal regression, <10% residual tumor), TRG 2 (partial regression, 10-50% residual tumor), and TRG 3 (minimal regression, >50% residual tumor).^15^ Two independent pathologists, blinded to treatment allocation, evaluated the specimens.

### Follow-up Strategy

#### Comprehensive long-term follow-up ensured complete data capture

Following the completion of primary treatment, patients entered long-term follow-up for survival endpoints. Follow-up visits were scheduled every 3 months for the first 2 years post-surgery, every 6 months for years 3-5, and annually thereafter. Vital status was verified through multiple sources: direct clinical follow-up, telephone contact, cross-referencing with local death registry records, and review of medical records from other healthcare facilities when applicable.

### Statistical Analysis

#### Protocol and Statistical Analysis Plan Documentation

The original study protocol (dated January 10, 2017) and statistical analysis plan outlined therein were submitted with this manuscript per NEJM requirements. The long-term survival analysis reported herein was pre-specified as a secondary endpoint in the original protocol, with overall survival defined as "time from randomization to death from any cause" and disease-free survival defined as "time from randomization to relapse of the disease." All survival analyses were performed according to the pre-specified statistical plan outlined in the original protocol, with no deviations from the planned analytical approach. The protocol document, including the detailed statistical plan and all study procedures, was submitted as part of the protocol documentation package.

### Study Design and Analysis Approach

The Dragon III trial was designed as an exploratory phase 2 study without formal power calculations for survival endpoints. The trial recruitment was completed after the surgical treatment of the 55th patient who met criteria for per-protocol analysis, consistent with the original protocol’s exploratory design and empirical sample size estimation. This stopping point was pre-planned to obtain preliminary data for efficacy assessment and to inform future phase III trial design, as outlined in the original protocol. All analyses were conducted according to the pre-specified statistical plan without knowledge of survival outcomes at the time of study completion.

### All analyses followed intention-to-treat principles with robust methodology

All survival analyses were performed according to the intention-to-treat principle, including all 74 randomized patients regardless of treatment completion or protocol adherence. Overall survival and disease-free survival were estimated using the Kaplan-Meier method. Survival differences between treatment groups were assessed using the log-rank test. Hazard ratios with 95% confidence intervals were estimated using Cox proportional hazards regression.

### Comprehensive subgroup and multivariable analyses were performed

Post-hoc subgroup analyses were performed for tumor regression grade, type of gastrectomy, postoperative pathological stage, and other baseline characteristics. Cox regression with stepwise selection was used to identify independent prognostic factors. All analyses were performed using SPSS version 30.0. Figures were created using R language version 2025. Two-sided p-values <0.05 were considered statistically significant. Two independent statisticians verified all analyses.

## RESULTS

### Patient Enrollment and Treatment Completion

Between August 2018 and March 2020, 74 patients with locally advanced gastric adenocarcinoma were randomized to neoadjuvant FLOT (n=40) or SOX (n=34). Patient flow through the trial has been previously published.^14^

### Treatment completion rates were comparable between groups

In the FLOT group, nine patients did not proceed to surgery: one withdrew consent, four refused surgery, three required early surgery due to acute bleeding, and one experienced a serious adverse event (acute cerebral infarction). In the SOX group, 10 patients did not undergo planned surgery: two withdrew consent, three refused surgery, one died from grade IV hematological toxicity followed by multiple organ failure, three violated protocol (two requested alternative chemotherapy and one was diagnosed with peritoneal metastases after allocation), and one experienced deep venous thrombosis detected during surgical evaluation. This resulted in 55 patients (31 FLOT, 24 SOX) completing all planned chemotherapy and undergoing surgical resection. Baseline characteristics showed no significant differences between treatment groups (Table 1).

### Follow-up and Adjuvant Treatment

#### Complete long-term follow-up was achieved for all patients

With data cutoff on May 8, 2025, median follow-up was 65.7 months (range: 0.97-80.5) for the intention-to-treat population. Complete vital status was obtained for all patients: 63.7 months (range: 8.4-80.5) in the FLOT group and 66.7 months (range: 0.97-80.5) in the SOX group. No patients were lost to follow-up (Supplementary Table 1).

### Post-surgical treatment was well-balanced between groups

Among the 55 patients who underwent surgical resection, the majority (52.7%) initiated adjuvant therapy within 5-8 weeks post-surgery. Most patients (85.5%) completed their planned adjuvant regimen, and 72.7% received at least three cycles. No significant differences in adjuvant therapy parameters were observed between treatment groups (all p>0.05), confirming the comparability of post-surgical treatment between the FLOT and SOX arms (Supplementary Tables 2 and 3).

### Both regimens demonstrated favorable safety profiles

Safety data for neoadjuvant chemotherapy were reported in our initial publication.^14^ Briefly, there was no significant difference in chemotherapy-related hematological or non-hematological adverse effects between groups. Nine events of grade 3-4 hematological toxicity were observed in the FLOT group versus five in the SOX group. Both regimens demonstrated favorable toxicity profiles, with notably lower rates of leukopenia and neutropenia than reported in the original FLOT4 trial. This was potentially influenced by more frequent granulocyte-colony stimulating factor (GCSF) use in the FLOT group (94.9%) compared to the SOX group (43.3%), though GCSF was only used for treatment rather than prophylaxis per protocol. All 55 patients in the per-protocol population completed the planned chemotherapy with full doses of their respective regimens.

### Primary Survival Outcomes: Equivalent Long-term Results

#### Both regimens achieved exceptional overall survival with no significant difference

Survival data are summarized in Table 2. At data cutoff, 41 deaths had occurred (21 [52.5%] in the FLOT group and 20 [58.8%] in the SOX group). No significant difference in overall survival was observed between treatment groups, with median OS of 61.5 months (95% CI: not reached) for FLOT versus 67.8 months (95% CI: 25.7-109.9) for SOX (hazard ratio [HR] 1.101, 95% CI: 0.595-2.036; p=0.759) (Figure 1). The 3-year overall survival rates were 62.5% (95% CI: 47.4-77.6) for FLOT and 55.9% (95% CI: 39.2-72.6) for SOX.

**Figure 1:**
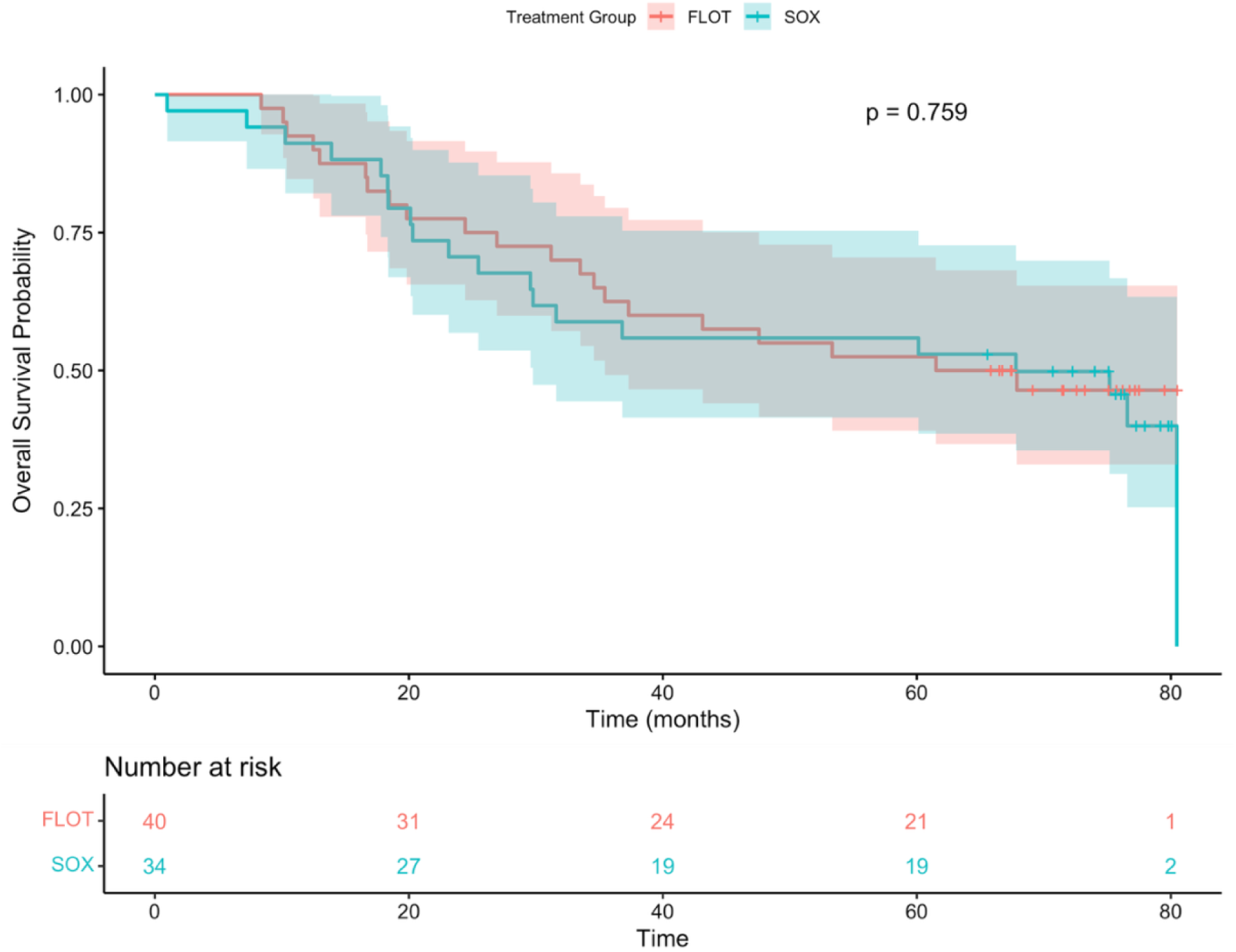
Overall Survival Kaplan-Meier Curves by Treatment Group. Kaplan-Meier estimates of overall survival for patients randomized to neoadjuvant FLOT (n=40) versus SOX (n=34) regimens. Shaded areas represent 95% confidence intervals. Numbers at risk are shown below the plot. Log-rank test p-value = 0.76 indicating no statistically significant difference between treatment groups.

**Table 2:** Overall Survival and Disease-Free Survival Outcomes.

| **Outcome** | **FLOT**<br>**(n=40)** | **Group SOX**<br>**(n=34)** | **Group Hazard Ratio (95% CI)** | **P-value** |
| --- | --- | --- | --- | --- |
| **Overall Survival** |  |  |  |  |
| Median OS, months (95% CI) | 61.5 (NR) | 67.8 (25.7-109.9) | 1.101 (0.595-2.036) | 0.759 |
| Deaths - no. (%) | 21 (52.5) | 20 (58.8) |  |  |
| 1-year OS rate, % (95% CI) | 90.0 (80.8-99.2) | 88.2 (77.4-99.0) |  |  |
| 2-year OS rate, % (95% CI) | 75.0 (61.7-88.3) | 67.6 (52.0-83.2) |  |  |
| 3-year OS rate, % (95% CI) | 62.5 (47.4-77.6) | 55.9 (39.2-72.6) |  |  |
| **Disease-Free Survival** |  |  |  |  |
| Median DFS, months (95% CI) | 23.0 (8.0-38.0) | 25.5 (12.0-38.9) | 1.060 (0.597-1.884) | 0.842 |
| Events - no. (%) | 25 (62.5) | 22 (64.7) |  |  |
| 1-year DFS rate, % (95% CI) | 67.3 (52.3-79.3) | 83.4 (67.4-92.6) |  |  |
| 2-year DFS rate, % (95% CI) | 43.5 (28.0-58.4) | 49.1 (31.4-65.1) |  |  |
| 3-year DFS rate, % (95% CI) | 31.6 (17.3-46.8) | 43.6 (26.4-59.8) |  |  |

**Abbreviations:** CI, confidence interval; DFS, disease-free survival; NR, not reached; OS, overall survival.

### Disease-free survival similarly demonstrated equivalent outcomes

Disease-free survival showed no statistically significant difference between groups (HR 1.060, 95% CI: 0.597-1.884; p=0.842), with median DFS of 23.0 months (95% CI: 8.0-38.0) for FLOT versus 25.5 months (95% CI: 12.0-38.9) for SOX (Figure 2). While 1-year DFS rates appeared numerically higher in the SOX group (83.4% vs 67.3%), this difference diminished at later time points, with 3-year DFS rates of 43.6% for SOX and 31.6% for FLOT.

**Figure 2:**
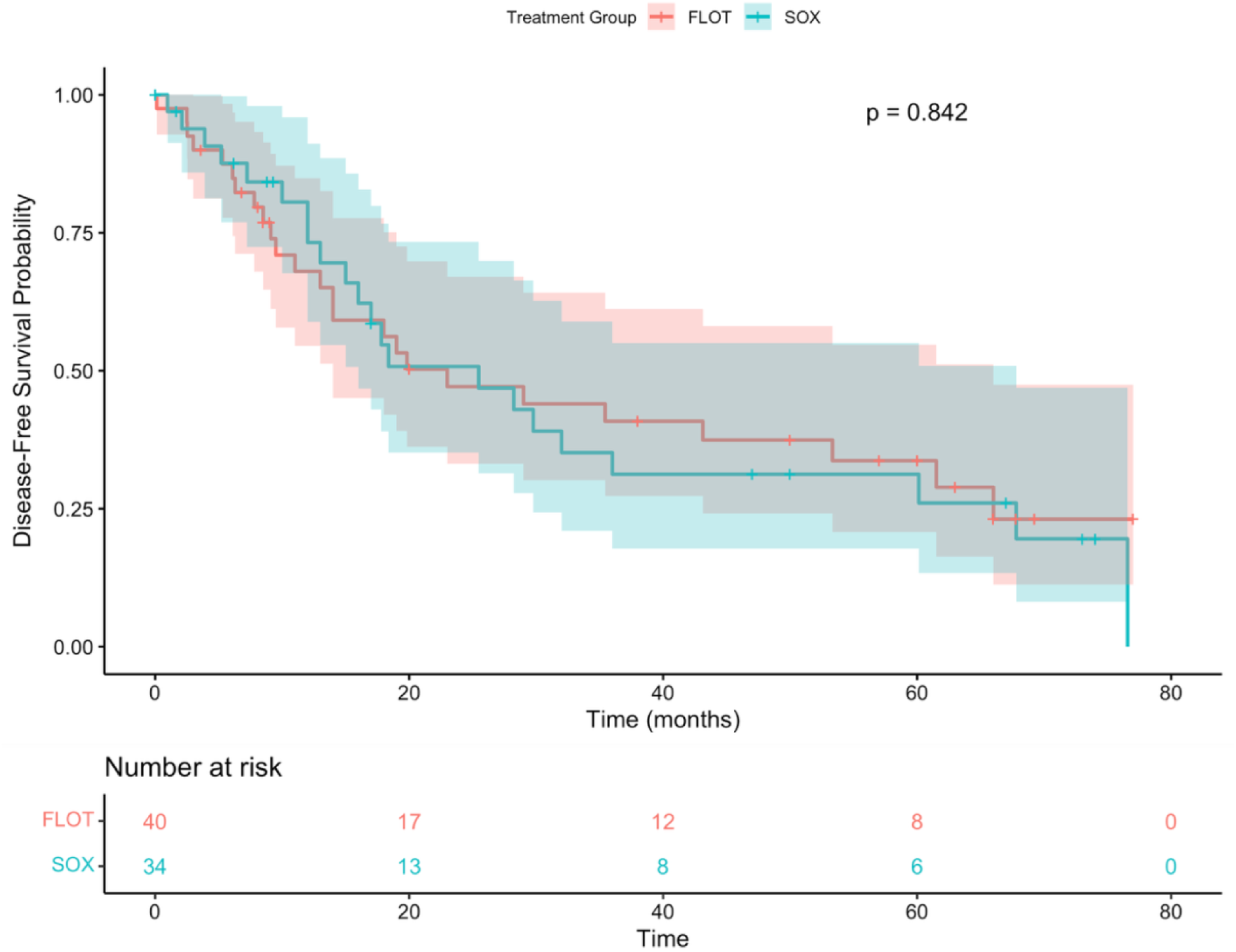
Disease-Free Survival Kaplan-Meier Curves by Treatment Group. Kaplan-Meier estimates of disease-free survival for patients randomized to neoadjuvant FLOT (n=40) versus SOX (n=34) regimens. Shaded areas represent 95% confidence intervals. Numbers at risk are shown below the plot. Log-rank test p-value = 0.84 indicating no statistically significant difference between treatment groups.

### Recurrence Patterns: Notable Differences in Metastatic Sites

#### Recurrence rates were similar between regimens

Recurrence sites are summarized in Table 3. Among 55 patients who underwent surgical resection, recurrence occurred in 27 (49.1%) patients during follow-up, with a numerically higher rate in the FLOT group (17/31, 54.8%) compared to the SOX group (10/24, 41.7%).

**Table 3:** Recurrence Sites by Treatment Group (Per-Protocol Population)

| Site of Recurrence | FLOT Group (n=31) | SOX Group (n=24) |
| --- | --- | --- |
|  | No. (%) | No. (%) |
| Any recurrence | 17 (54·8) | 10 (41·7) |
| Retroperitoneal lymph nodes | 7 (22·6) | 5 (20·8) |
| Peritoneal | 5 (16·1) | 4 (16·7) |
| Liver | 4 (12·9) | 2 (8·3) |
| Lung | 4 (12·9) | 0 (0·0) |
| Locoregional (anastomosis) | 3 (9·7) | 1 (4·2) |
| Bone | 1 (3·2) | 2 (8·3) |
| Brain | 1 (3·2) | 2 (8·3) |

FLOT=docetaxel, oxaliplatin, leucovorin, 5-fluorouracil; SOX=S-1 plus oxaliplatin. Per-protocol population includes only patients who underwent surgery.

### Most common recurrence sites were consistent between groups

The most common sites of recurrence in both treatment groups were retroperitoneal lymph nodes (FLOT: 22.6%; SOX: 20.8%) and peritoneum (FLOT: 16.1%; SOX: 16.7%). **Notably, lung metastases were observed** exclusively in the FLOT group (12.9% vs 0%), while brain metastases were more common in the SOX group (8.3% vs 3.2%).

### Prognostic Factors: Clinicopathological Features Outweigh Regimen Choice

#### Multiple clinicopathological factors significantly impacted survival outcomes

Comprehensive subgroup analyses for overall survival are summarized in Supplementary Table 4 and illustrated in Figure 3. Multiple factors were significantly associated with survival outcomes, with **pathological response proving more important than regimen selection.**

**Figure 3:**
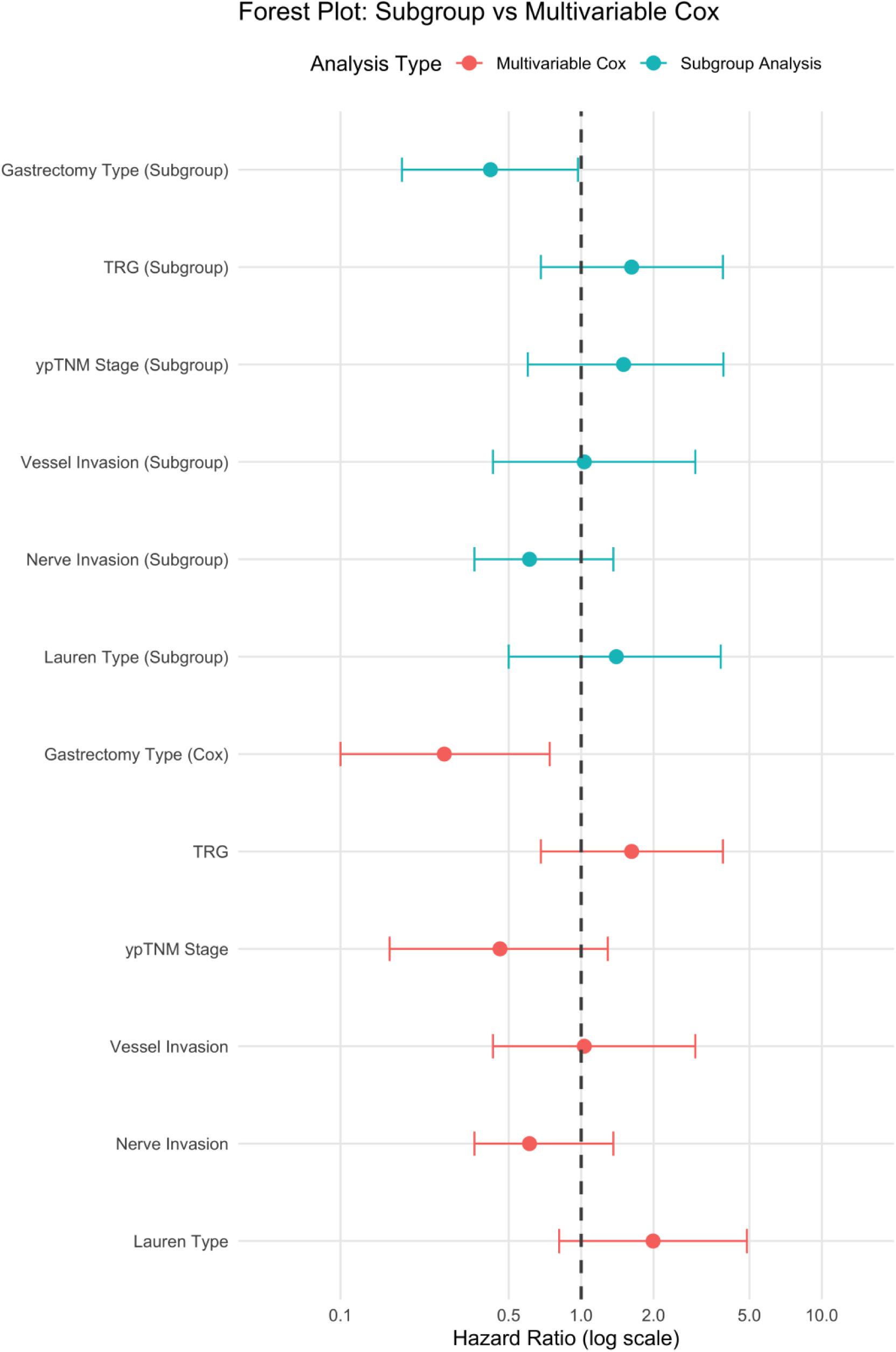
Forest Plot of Prognostic Factors for Overall Survival. Comparison of hazard ratios from subgroup analysis (blue circles) and multivariable Cox regression (red circles) for various prognostic factors. Hazard ratios are plotted on a logarithmic scale with 95% confidence intervals. The vertical dashed line at HR=1.0 represents no effect. Only gastrectomy type remained statistically significant in the multivariable model.

### Pathological response was strongly prognostic

Pathological response, as measured by tumor regression grade, showed significant impact on survival (p=0.017), with complete/subtotal regression (TRG 1a+1b) associated with markedly longer median OS (80.5 months) compared to partial/minimal regression (47.6 months).

### Surgical factors were equally critical

Gastrectomy extent emerged as another significant prognostic factor (p=0.003), with partial gastrectomy demonstrating superior outcomes compared to total gastrectomy (median OS not reached vs 37.3 months; HR 3.619, 95% CI: 1.360-9.628).

### Traditional prognostic markers maintained their significance

Pathological staging also influenced survival (p=0.025), with gradually decreasing survival from Stage I (median not reached) to Stage II (80.5 months) to Stage III (33.5 months). Both vessel invasion (p=0.021) and nerve invasion (p=0.041) were significant negative prognostic indicators. Lauren classification demonstrated prognostic value (p=0.023), with intestinal type showing more favorable outcomes compared to other histological subtypes. **Across all prognostic factors, patients who dropped out before surgery consistently showed the poorest outcomes (median OS 29.6 months).**

### Independent Prognostic Factors: Surgery Emerges as Key Driver

#### Gastrectomy type was the only independent survival predictor

Cox regression with stepwise selection identified gastrectomy type as the only independent predictor of overall survival (p=0.002) (Table 4, Figure 3). Compared to partial gastrectomy: total gastrectomy HR 3.619 (95% CI: 1.360-9.628; p=0.010), unknown gastrectomy HR 5.134 (95% CI: 1.844-14.291; p=0.002).

**Table 4:** Multivariable Analysis of Overall Survival.

| Variable | Hazard Ratio (95% CI) | p-value |
| --- | --- | --- |
| Type of Gastrectomy |  | 0·002 |
| Partial gastrectomy | Reference |  |
| Total gastrectomy | 3·619 (1·360-9·628) | 0·010 |
| Dropped cases | 5·134 (1·844-14·291) | 0·002 |

CI=confidence interval.
Variables initially considered but removed during stepwise selection: treatment group (FLOT vs SOX), tumour regression grade, Lauren classification, and postoperative pathological stage (ypTNM).
FLOT=docetaxel, oxaliplatin, leucovorin, 5-fluorouracil; SOX=S-1 plus oxaliplatin; ypTNM=postoperative pathological TNM stage.

**Importantly, treatment group, tumor regression grade, Lauren classification, and postoperative stage were eliminated during stepwise selection,** reinforcing that surgical approach—not regimen choice—was the primary modifiable factor influencing long-term survival.

## DISCUSSION

This first prospective head-to-head comparison definitively establishes equivalent long-term survival between neoadjuvant FLOT and SOX regimens, fundamentally changing treatment selection for locally advanced gastric cancer. With both regimens achieving exceptional median overall survival exceeding 5 years (61.5 vs 67.8 months, p=0.759), clinicians worldwide now have evidence-based freedom to select regimens based on patient and institutional factors rather than survival concerns.

### Clinical Context and Validation

Our survival results align with the FLOT4 trial’s median overall survival of 50 months,^7^ though direct comparisons require caution given differences in patient populations and staging systems. Despite lower pathological response rates with FLOT in our Asian population (20.0% vs 37% in FLOT4), long-term survival outcomes remained excellent, suggesting pathological response may not fully capture clinical benefit in all patients.^6^ This finding is consistent with previous studies suggesting that while tumor regression grade is prognostic, the relationship between pathological response and survival may vary across populations and regimens.^16^

### The Primacy of Clinicopathological Factors

The study’s most practice-changing insight is that several clinicopathological factors were stronger survival predictors than neoadjuvant regimen choice. This paradigm shift redirects clinical focus from "which regimen is better" to "how do we optimize response and surgical management."

### Pathological response emerged as a critical predictor

Tumor regression grade, gastrectomy type, vessel invasion, nerve invasion, and Lauren classification^17^ all demonstrated prognostic significance in our analysis. Patients achieving complete/subtotal tumor regression had markedly better survival (80.5 vs 47.6 vs 29.6 months for TRG 1a+1b vs 2+3 vs dropout, p=0.017), reinforcing the importance of pathological response as a prognostic factor. This aligns with foundational work by Becker et al. and Mandard et al., who established tumor regression grading systems that remain essential for prognostic assessment in upper gastrointestinal cancers.^15,18^

### Surgical factors proved equally critical

Gastrectomy type emerged as the strongest independent survival predictor (HR 3.619 for total vs partial gastrectomy, p=0.010). While this may reflect tumor location and disease extent, it highlights surgical factors’ impact on outcomes. Superior outcomes with partial gastrectomy may relate to preserved function, reduced morbidity, and selection of patients with more favorable characteristics. This finding is consistent with contemporary surgical series demonstrating that extent of resection influences both immediate and long-term outcomes in gastric cancer patients.

### Traditional prognostic factors maintained their relevance

The observed associations between vessel invasion, nerve invasion, and survival further strengthen our prognostic model. Patients with negative vessel invasion demonstrated significantly better survival (80.5 vs 31.2 months, p=0.021) compared to those with positive findings. Similarly, negative nerve invasion was associated with superior outcomes (not reached vs 47.6 months, p=0.041). Lauren’s classification^17^ also demonstrated prognostic significance, with intestinal-type histology associated with better survival than other histological types (not reached vs 47.6 months, p=0.023).

### Global Treatment Implications

The equivalent outcomes between FLOT and SOX support findings from recent Asian trials, including RESOLVE and RESONANCE, which demonstrated promising efficacy for SOX regimens.^10,11^ These results, along with findings from other major Asian gastric cancer trials,^19^ reinforce the importance of optimizing treatment selection for Asian populations. Our study provides the first prospective head-to-head survival comparison between these regimens, supporting either approach based on institutional and patient factors. The demonstrated equivalence has important implications for treatment selection in Asian populations, where SOX offers practical advantages including oral administration and reduced treatment cycles.

Disease-free survival showed no significant difference between groups (23.0 vs 25.5 months, p=0.842). Sites of recurrence differed notably, with no lung metastases in the SOX group versus 12.9% in the FLOT group, warranting validation in larger studies. These differences may reflect distinct drug-specific effects on sites of metastases or could be attributed to the relatively small sample size.

The high adjuvant therapy completion rate (85.5%) and similar treatment adherence indicate both regimens were well-tolerated and feasible in the perioperative setting. This is particularly important given that completion of planned perioperative therapy is associated with improved outcomes in gastric cancer patients.^7^

### Future Treatment Landscape and Regimen Selection

Given equivalent survival outcomes, regimen selection should be individualized. SOX offers practical advantages including reduced cycles (3 vs 4), fewer hospital visits, and potentially lower neutropenia rates.^14^ Patient factors (age, performance status, treatment preference) and institutional considerations (experience, protocols, cost) should guide decision-making.

Our findings must be interpreted within the evolving landscape of gastric cancer treatment. Recent advances in targeted therapy and immunotherapy are transforming treatment paradigms.^20,21^ The integration of trastuzumab deruxtecan for HER2-positive disease and immune checkpoint inhibitors represents significant progress.^21,22^ The DRAGON IV trial’s demonstration of improved pathological response with immunotherapy plus chemotherapy suggests these combinations represent the next treatment frontier.^13^ Future research should explore whether the survival equivalence we observed between FLOT and SOX is maintained when combined with immunotherapy and targeted agents.

### Study Limitations and Clinical Integration

Study limitations include the phase 2 design underpowered for survival endpoints, single-center setting limiting generalizability, and exclusively Asian population. The observed hazard ratios near 1.0 suggest true equivalence, but phase 3 trials would be needed for definitive non-inferiority demonstration.

For immediate clinical application, these results support individualized treatment selection based on practical considerations rather than survival expectations. The key clinical message is clear: achieving optimal pathological response and performing appropriate surgical resection are more critical than the specific neoadjuvant regimen chosen. This insight should guide treatment planning, with emphasis on maximizing response rates and ensuring access to expert surgical management.

### Interpretation

This definitive 5-year analysis resolves a fundamental question in gastric cancer treatment: FLOT and SOX neoadjuvant regimens deliver equivalent excellent survival outcomes, both achieving median overall survival exceeding 5 years. This finding transforms treatment selection from survival-based concerns to decisions guided by patient preferences, institutional expertise, and practical considerations.

The study’s most practice-changing insight is that optimizing pathological response and surgical approach—rather than regimen choice—represents the true opportunity to improve outcomes. Patients achieving complete or subtotal tumor regression demonstrated 80.5-month median survival, while gastrectomy type emerged as the strongest independent predictor. These findings redirect clinical focus toward maximizing treatment response and surgical excellence, regardless of chosen regimen.

For global practice, these results provide definitive guidance: clinicians can confidently offer either FLOT or SOX based on individualized factors without compromising survival expectations. This is particularly significant for Asian populations, where SOX offers practical advantages—oral administration and fewer cycles—while maintaining equivalent outcomes to the established FLOT standard.

Both regimens now serve as validated platforms for integration with emerging immunotherapy and targeted agents. The demonstrated equivalence creates opportunities for personalized approaches and biomarker-driven selection strategies in future combination trials.

These findings represent rational treatment selection in modern oncology: achieving excellent outcomes through multiple effective approaches while focusing on what truly drives survival— optimal pathological response and expert surgical management. This study establishes the foundation for personalized perioperative gastric cancer care.

## Data Availability

Individual patient data supporting the conclusions will be made available to qualified investigators upon reasonable request, subject to appropriate data-sharing agreements, and ethics approval.

https://clinicaltrials.gov/study/NCT03636893

## Contributors

BKS conceived and designed the study, collected the patient data, performed the data analysis, and drafted the manuscript. CL and ZGZ designed the study and edited the final manuscript. All authors met the criteria for authorship, according to the guidelines of the International Committee of Medical Journal Editors.

## Acknowledgments

We thank the patients and their families for participating in this trial. We acknowledge the contributions of the nursing staff, research coordinators, and healthcare professionals involved in patient care and data collection. We thank the Department of Pathology, Department of Medical Oncology, Department of Radiology, and Clinical Research Center at Ruijin Hospital for their collaboration and support throughout this study.

## Trial Registration

ClinicalTrials.gov identifier: NCT03636893

## SUPPLEMENTARY TABLES

**Supplementary Table 1:**
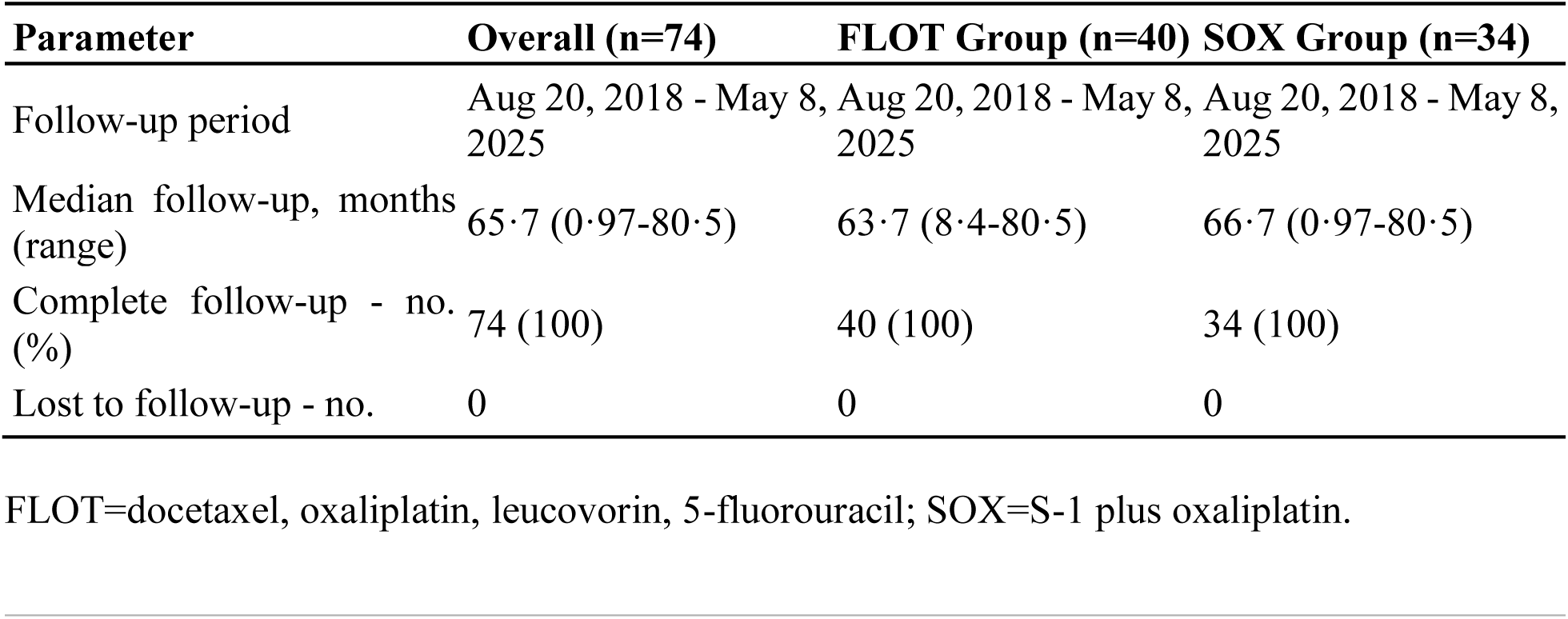
Follow-up Summary.

**Supplementary Table 2:** Adjuvant Chemotherapy Completion (n=55)

| **Variable** | **Category** | **Frequency** | **Percent** |
| --- | --- | --- | --- |
| **Adjuvant Chemo Start Timing** |  |  |  |
|  | ≤4 weeks | 7 | 12.7% |
|  | 5-8 weeks | 29 | 52.7% |
|  | ≥9 weeks | 9 | 16.4% |
|  | 2nd line chemo | 10 | 18.2% |
| **Second-line Chemotherapy** |  |  |  |
|  | No | 37 | 67.3% |
|  | Yes | 10 | 18.2% |
|  | Unknown | 8 | 14.5% |
| **Adjuvant Chemo Cycles** |  |  |  |
|  | ≥3 cycles | 40 | 72.7% |
|  | 1-2 cycles | 7 | 12.7% |
|  | Unknown | 8 | 14.5% |
| **Per Protocol Chemo** |  |  |  |
|  | Yes | 34 | 61.8% |
|  | No | 13 | 23.6% |
|  | Unknown | 8 | 14.5% |
| **Adjuvant Chemo Completion** |  |  |  |
|  | Yes | 47 | 85.5% |
|  | Unknown | 8 | 14.5% |
| **Performance Status at last follow up** |  |  |  |
|  | Good | 39 | 70.9% |
|  | Poor | 15 | 27.3% |
|  | Unknown | 1 | 1.8% |
| **Post-operative Follow up at Ruijin** |  |  |  |
|  | Yes | 48 | 87.3% |
|  | No | 7 | 12.7% |

Chemo=chemotherapy.

**Supplementary Table 3:** Comparison of Adjuvant Chemotherapy.

| Variable | Category | FLOT<br>(n=31) | SOX<br>(n=24) | Total<br>(n=55) | p-value* |
| --- | --- | --- | --- | --- | --- |
| **Adjuvant Chemo Start Timing** |  |  |  |  | 0·673 |
|  | ≤4 weeks | 4 (12·9%) | 3 (12·5%) | 7 (12·7%) |  |
|  | 5-8 weeks | 17 (54·8%) | 12 (50·0%) | 29 (52·7%) |  |
|  | ≥9 weeks | 6 (19·4%) | 3 (12·5%) | 9 (16·4%) |  |
|  | 2nd line<br>chemo | 4 (12·9%) | 6 (25·0%) | 10 (18·2%) |  |
| **Second-line Chemotherapy** |  |  |  |  | 0·508 |
|  | No | 22 (71·0%) | 15 (62·5%) | 37 (67·3%) |  |
|  | Yes | 6 (19·4%) | 4 (16·7%) | 10 (18·2%) |  |
|  | Unknown | 3 (9·7%) | 5 (20·8%) | 8 (14·5%) |  |
| **Adjuvant Chemo Cycles** |  |  |  |  | 0·318 |
|  | ≥3 cycles | 25 (80·6%) | 15 (62·5%) | 40 (72·7%) |  |
|  | 1-2 cycles | 3 (9·7%) | 4 (16·7%) | 7 (12·7%) |  |
|  | Unknown | 3 (9·7%) | 5 (20·8%) | 8 (14·5%) |  |
| **Per Protocol Adjuvant Chemo** |  |  |  |  | 0·501 |
|  | Yes | 20 (64·5%) | 14 (58·3%) | 34 (61·8%) |  |
|  | No | 8 (25·8%) | 5 (20·8%) | 13 (23·6%) |  |
|  | Unknown | 3 (9·7%) | 5 (20·8%) | 8 (14·5%) |  |
| **Adjuvant Chemo Completion** |  |  |  |  | 0·245 |
|  | Yes | 28 (90·3%) | 19 (79·2%) | 47 (85·5%) |  |
|  | Unknown | 3 (9·7%) | 5 (20·8%) | 8 (14·5%) |  |
| **Performance Status at last follow up** |  |  |  |  | 0·483 |
|  | Good | 23 (74·2%) | 16 (66·7%) | 39 (70·9%) |  |
|  | Poor | 8 (25·8%) | 7 (29·2%) | 15 (27·3%) |  |
|  | Unknown | 0 (0·0%) | 1 (4·2%) | 1 (1·8%) |  |
| **Post-operative Follow up at Ruijin** |  |  |  |  | 0·441 |
|  | Yes | 28 (90·3%) | 20 (83·3%) | 48 (87·3%) |  |
|  | No | 3 (9·7%) | 4 (16·7%) | 7 (12·7%) |  |

\*Chi-square test or Fisher's exact test where appropriate. Chemo=chemotherapy; FLOT=docetaxel, oxaliplatin, leucovorin, 5-fluorouracil; SOX=S-1 plus oxaliplatin.

**Supplementary Table 4:**
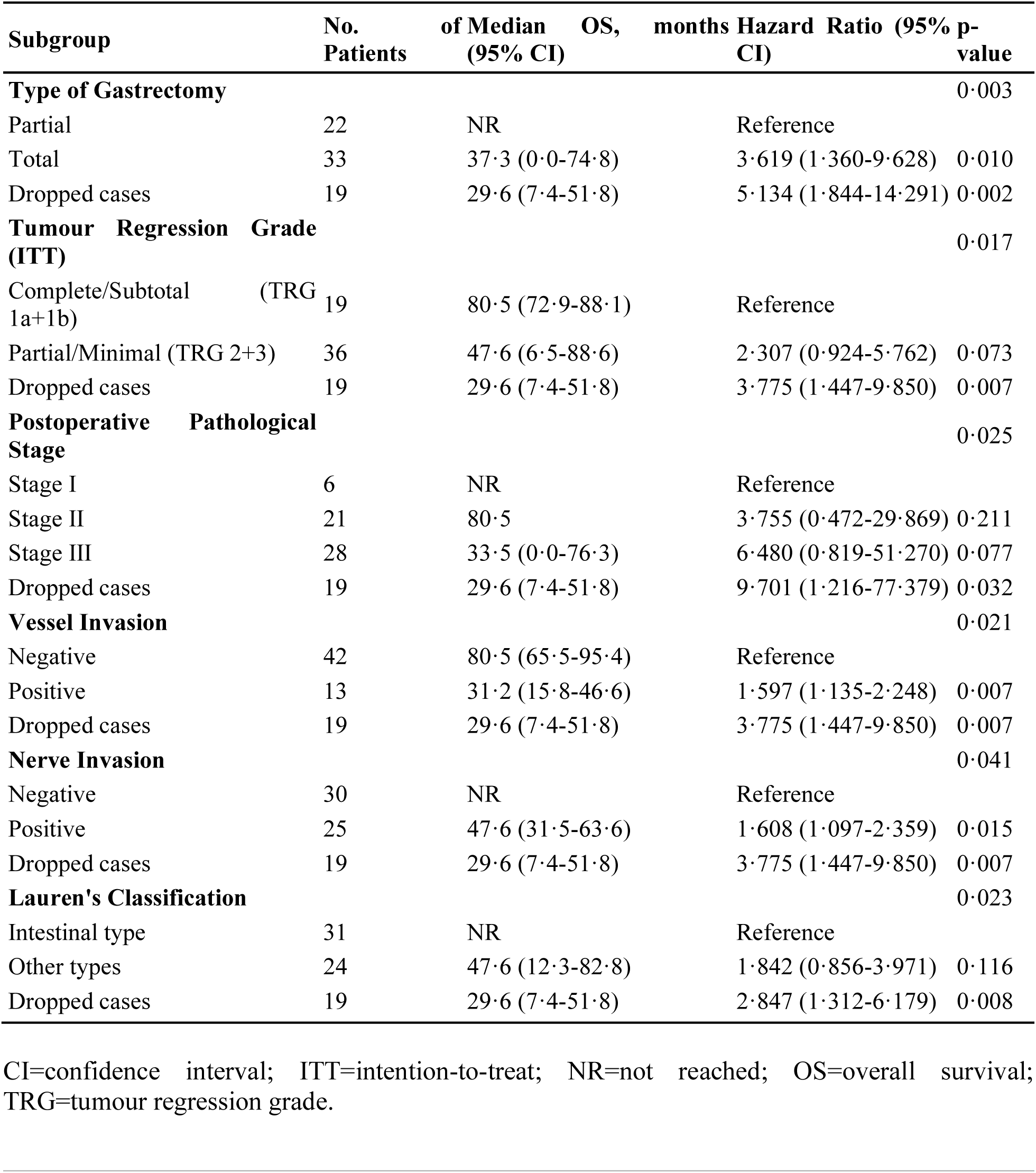
Subgroup Analysis of Overall Survival (n=74)

## REFERENCES

1. Sung H, Ferlay J, Siegel RL, et al. Global Cancer Statistics 2020: GLOBOCAN estimates of incidence and mortality worldwide for 36 cancers in 185 countries. CA Cancer J Clin 2021; 71: 209–49.

2. Cunningham D, Allum WH, Stenning SP, et al. Perioperative chemotherapy versus surgery alone for resectable gastroesophageal cancer. N Engl J Med 2006; 355: 11–20.

3. Ronellenfitsch U, Schwarzbach M, Hofheinz R, et al. Perioperative chemo(radio)therapy versus primary surgery for resectable adenocarcinoma of the stomach, gastroesophageal junction, and lower esophagus. Cochrane Database Syst Rev 2013; 2013: CD008107.

4. Ychou M, Boige V, Pignon JP, et al. Perioperative chemotherapy compared with surgery alone for resectable gastroesophageal adenocarcinoma: an FNCLCC and FFCD multicenter phase III trial. J Clin Oncol 2011; 29: 1715–21.

5. Bang YJ, Kim YW, Yang HK, et al. Adjuvant capecitabine and oxaliplatin for gastric cancer after D2 gastrectomy (CLASSIC): a phase 3 open-label, randomised controlled trial. Lancet 2012; 379: 315–21.

6. Al-Batran SE, Hofheinz RD, Pauligk C, et al. Histopathological regression after neoadjuvant docetaxel, oxaliplatin, fluorouracil, and leucovorin versus epirubicin, cisplatin, and fluorouracil or capecitabine in patients with resectable gastric or gastro-oesophageal junction adenocarcinoma (FLOT4-AIO): results from the phase 2 part of a multicentre, open-label, randomised phase 2/3 trial. Lancet Oncol 2016; 17: 1697–708.

7. Al-Batran SE, Homann N, Pauligk C, et al. Perioperative chemotherapy with fluorouracil plus leucovorin, oxaliplatin, and docetaxel versus fluorouracil or capecitabine plus cisplatin and epirubicin for locally advanced, resectable gastric or gastro-oesophageal junction adenocarcinoma (FLOT4): a randomised, phase 2/3 trial. Lancet 2019; 393: 1948–57.

8. Wagner AD, Syn NL, Moehler M, et al. Chemotherapy for advanced gastric cancer. Cochrane Database Syst Rev 2017; 8: CD004064.

9. Smyth EC, Nilsson M, Grabsch HI, van Grieken NC, Lordick F. Gastric cancer. Lancet 2020; 396: 635–48.

10. Zhang X, Liang H, Li Z, et al. Perioperative or postoperative adjuvant oxaliplatin with S- 1 versus adjuvant oxaliplatin with capecitabine in patients with locally advanced gastric or gastro-oesophageal junction adenocarcinoma undergoing D2 gastrectomy (RESOLVE): final report of a randomised, open-label, phase 3 trial. Lancet Oncol 2025; 26: 312–9.

11. Wang X, Lu C, Wei B, et al. Perioperative versus adjuvant S-1 plus oxaliplatin chemotherapy for stage II/III resectable gastric cancer (RESONANCE): a randomized, open-label, phase 3 trial. J Hematol Oncol 2024; 17: 17.

12. Yamada Y, Higuchi K, Nishikawa K, et al. Phase III study comparing oxaliplatin plus S-1 with cisplatin plus S-1 in chemotherapy-naïve patients with advanced gastric cancer. Ann Oncol 2015; 26: 141–8.

13. Li C, Tian Y, Zheng Y, et al. Pathologic Response of Phase III Study: Perioperative Camrelizumab Plus Rivoceranib and Chemotherapy Versus Chemotherapy for Locally Advanced Gastric Cancer (DRAGON IV/CAP 05). J Clin Oncol 2025; 43: 464–74.

14. Sah BK, Zhang B, Zhang H, et al. Neoadjuvant FLOT versus SOX phase II randomized clinical trial for patients with locally advanced gastric cancer. Nat Commun 2020; 11: 6093.

15. Becker K, Mueller JD, Schulmacher C, et al. Histomorphology and grading of regression in gastric carcinoma treated with neoadjuvant chemotherapy. Cancer 2003; 98: 1521–30.

16. Tsagkalidis V, Blaszczyk MB, In H. Interpretation of Tumor Response Grade following Preoperative Therapy for Gastric Cancer: An Overview. Cancers (Basel) 2023; 15: 2847.

17. Lauren P. The two histological main types of gastric carcinoma: diffuse and so-called intestinal-type carcinoma. An attempt at a histo-clinical classification. Acta Pathol Microbiol Scand 1965; 64: 31–49.

18. Mandard AM, Dalibard F, Mandard JC, et al. Pathologic assessment of tumor regression after preoperative chemoradiotherapy of esophageal carcinoma. Clinicopathologic correlations. Cancer 1994; 73: 2680–86.

19. Lee J, Lim DH, Kim S, et al. Phase III trial comparing capecitabine plus cisplatin versus capecitabine plus cisplatin with concurrent capecitabine radiotherapy in completely resected gastric cancer with D2 lymph node dissection: the ARTIST trial. J Clin Oncol 2012; 30: 268–73.

20. Joshi SS, Badgwell BD. Current treatment and recent progress in gastric cancer. CA Cancer J Clin 2021; 71: 264–79.

21. Shitara K, Bang YJ, Iwasa S, et al. Trastuzumab deruxtecan in previously treated HER2- positive gastric cancer. N Engl J Med 2020; 382: 2419–30.

22. Kang YK, Boku N, Satoh T, et al. Nivolumab in patients with advanced gastric or gastro-oesophageal junction cancer refractory to, or intolerant of, at least two previous chemotherapy regimens (ONO-4538-12, ATTRACTION-2): a randomised, double-blind, placebo-controlled, phase 3 trial. Lancet 2017; 390: 2461-71.

